# Recent advances in the clinical applications of machine learning in proton therapy

**DOI:** 10.1101/2024.10.09.24314920

**Authors:** Vanessa L. Wildman, Jacob F. Wynne, Aparna H. Kesarwala, Xiaofeng Yang

## Abstract

The present systematic review is an effort to explore the different clinical applications and current implementations of machine/deep learning in proton therapy. It will assist as a reference for scientists, researchers, and other health professionals who are working in the field of proton radiation therapy and need up-to-date knowledge regarding recent technological advances. This review utilized Pubmed and Embase to search for and identify research studies of interest published between 2019 and 2024. This systematic literature review utilized PubMed and Embase to search for and identify studies pertinent to machine learning in proton therapy. The time period of 2019 to 2024 was chosen to capture the most recent signficant advances. An initial search on PubMed was made with the search strategy “‘proton therapy’, ‘machine learning’, ‘deep learning’”, with filters including only research articles from 2019 to 2024, returning 84 results. Next, “(“proton therapy”) AND (“machine learning” OR “deep learning”)” was searched on Embase, retrieving 546 results. When filtered between 2019 to 2024 and to only research articles, 250 results were retrieved on Embase. Reviews, editorials, technical notes, and articles in any language other than English were excluded from the broad search on both databases. Filtering by title, papers were chosen based on two inclusion factors: explicit application to, or mention of, proton therapy, and inclusion of a machine learning algorithm. Assessing by abstract, works irrelevant to specific aspects of the proton therapy workflow in the scope of the review were excluded. Upon assessing and evaluating full texts for quality, studies were excluded that lacked a clear explanation of model architecture. If multiple studies of the same architecture applied to the same workflow step were identified, chronologically only the most recent advancement in application was included. An additional 5 studies that met all inclusion criteria were identified from references of chosen papers. In total, 38 relevant studies have been summarized and incorporated into this review. This is the first systematic review to comprehensively cover all current and potential areas of application of machine learning to the proton therapy clinical workflow.

## Introduction

The usage of protons in radiation was first proposed in 1946 by Wilson, a Harvard particle physicist [1]. The first treatments on human patients began in 1954 at the Berkeley Radiation Laboratory [2], and the first therapy center was opened in Boston in 1961. In 1988, the FDA approved proton therapy for clinical use. Currently, the National Association for Proton Therapy (NAPT) reports 45 active proton therapy centers located across the United States, and a recent clinical investigation found that between 2012 and 2021, the total annual number of patients treated with proton therapy increased from 5,377 to 15,829 [3]. Demand for proton therapy continues to rise, due to its increased precision and reduction of integral dose.

The physics behind proton therapy originates from the unique energetic nature of the proton. As beams travel through tissues, protons undergo inelastic and elastic Coulombic interactions. Kinetic energy is continuously lost through frequent inelastic interactions with atomic electrons. However, most protons continue to travel in a nearly straight line through the medium as their rest mass is significantly larger than that of an electron and only the electron is ejected. A proton passing close to the nucleus experiences a repulsive elastic interaction which, owing to the large mass of the nucleus, deflects the proton from its originally straight trajectory [4]. Such interactions are less frequent, but must be considered in treatment planning as they result in a change of particle trajectory. As energy loss of the proton beam determines range in the patient, an understanding of these interactions in the clinical context is essential. The Bragg peak is the fundamental phenomena supporting the efficacy of proton therapy treatment. The Bragg curve plots the energy loss of ionizing radiation, such as photons or protons, during its travel through matter. For protons, the peak occurs immediately prior to the particles coming to rest. This is the depth where the proton will deposit most of its dose [4]. With precise planning, this unique property has the potential to spare skin and other healthy tissue from radiation exposure both proximal and distal to the tumor target.

Conventional photon therapy and proton therapy each possess unique advantages and disadvantages. Because they are uncharged particles, photons primarily interact with matter through indirect processes such as Compton scattering and the photoelectric effect. These interactions result in a relatively uniform deposition of energy along their path through tissue, which leads to broader scatter to surrounding tissue and skin in the setting of conventional radiotherapy. Photon therapy is associated with resultant side effects, such as radiation dermatitis, due to increased skin exposure [5]. As a result of the Bragg peak, proton radiation is delivered with greater precision to the tumor site, sparing healthy surrounding tissue and skin relative to conventional photon radiotherapy. Specific techniques such as pencil beam scanning and intensity modulated proton therapy have been developed to further optimize proton therapy treatment delivery. Proton therapy is currently preferred in pediatric cases and in scenarios where tumor location is adjacent to sensitive organs at risk (OARs), such as in cancers of the head and neck and central nervous system. It is also useful in geometrically complex treatments with challenging dosimetry, such as when treating synchronous bilateral breast cancers.

Numerous machine learning (ML) applications are currently employed in conventional radiotherapy and may easily be adapted to proton therapy [6]. At present, models are employed primarily in segmentation, dose calculation and risk management. ML methods to improve clinical decision-making are needed. Adapting existing ML protocols for traditional photon treatment to proton applications may be expected to improve these processes, enhancing physician efficiency and quality in treatment planning and delivery.

ML, as a subfield of artificial intelligence (AI), encompasses models which learn from datasets to generate decisions without being explicitly coded for a repetitive, standard task. ML models may be trained using supervised on unsupervised techniques. Supervised learning provides the model labeled and correctly paired training and test datasets, where the correct output is known for each input [7]. These methods are typically applied to classification tasks where the goal is to map outputs from a given input, such as verifying Bragg peak range or optimizing treatment plans with Knowledge-Based Planning (KBP). In contrast, unsupervised learning is commonly applied to complex tasks where labeled datasets are not readily available, such as imaging segmentation or contouring. Deep learning (DL) is a subset of ML which utilizes neural networks. Inspired by information processing in the brain, neural networks pass raw input data through layers of nodes, each comprised of simple mathematical operations, to create an output. With additional (deeper) layers, more complex functions, such as image recognition, are possible [8]. Convolutional neural networks (CNNs) are specialized DL models equipped to process grid-like data. This is beneficial in medical imaging, as CNNs can extract features from the pixels or voxels of patient imaging data across several modalities. The layers of a CNN, known as convolutional layers, apply kernels to the input image to extract specific features such as contours or patterns across progressively smaller spatial scales [9].

A common form of DL that has been applied in the planning stage of radiotherapy treatment delivery is the Generative Adversarial Network (GAN). This type of network was originally created in 2017 by Goodfellow *et al*. and is a novel approach to unpaired image-to-image translation superior to traditional DL models when generating synthetic images [10]. The GAN architecture consists of two connected neural networks, a generator which creates synthetic data, and a discriminator which evaluates the resemblance to the real data. These two neural networks are trained simultaneously, using an adversarial framework that ultimately guides the generator to improve performance with each iteration, producing increasingly realistic data that challenges the discriminator. Training is complete when the min-max loss function is optimized. GANs are particularly useful in complex image translation tasks, such as converting images from one modality to another (e.g. CT to MRI). U-Nets, CNNs with a U-shaped architecture, are particularly suited to tasks in image segmentation, due to their ability to extract and retain image details across several spatial scales [11]. Such a network is able to be trained on few images, and performs segmentation-related tasks very quickly.

## Methods

This review utilized Pubmed and Embase to search for and identify research studies of interest published between 2019 and 2024. This systematic literature review utilized PubMed and Embase to search for and identify studies pertinent to machine learning in proton therapy. The time period of 2019 to 2024 was chosen to capture the most recent signficant advances. An initial search on PubMed was made with the search strategy “‘proton therapy’, ‘machine learning’, ‘deep learning’”, with filters including only research articles from 2019 to 2024, returning 84 results. Next, “(“proton therapy”) AND (“machine learning” OR “deep learning”)” was searched on Embase, retrieving 546 results. When filtered between 2019 to 2024 and to only research articles, 250 results were retrieved on Embase. Reviews, editorials, technical notes, and articles in any language other than English were excluded from the broad search on both databases. Filtering by title, papers were chosen based on two inclusion factors: explicit application to, or mention of, proton therapy, and inclusion of a machine learning algorithm. Assessing by abstract, works irrelevant to specific aspects of the proton therapy workflow in the scope of the review were excluded. Upon assessing and evaluating full texts for quality, studies were excluded that lacked a clear explanation of model architecture. If multiple studies of the same architecture applied to the same workflow step were identified, chronologically only the most recent advancement in application was included. An additional 5 studies that met all inclusion criteria were identified from references of chosen papers. In total, 38 relevant studies have been summarized and incorporated into this review.

## Results

### 1. ML for Patient Selection and Predictive Outcome Modeling in Proton Therapy

ML models are being developed to improve the efficiency of pre-screening processes and determining which modality of radiotherapy will result in the least adverse effects. Pre-selection enables patient informatics to be compared to an established criteria protocol that assesses whether or not an individual is eligible for, or will respond better to, proton therapy versus photon radiotherapy. This automated method enables more patients to be pre-screened in a shorter amount of time as well as eliminating any physician bias in the selection process. It is also possible to utilize a predictive model to choose proton or photon therapy based on post-radiation toxicity prediction as a marker of preferability.

#### 1.1 Pre-screening for Therapy Selection

Kouwenberg *et al*. [12] developed a pre-selection algorithm specifically designed for intensity modulated proton therapy (IMPT) for head and neck cancer patients. The group found that the proposed algorithm significantly reduces time and labor costs by utilizing ML and automated planning. The algorithm created a fully-automated IMPT plan for patients with a prior model-based selection (MBS) approach for photon treatment. A Gaussian naive Bayes classifier for MBS outcome prediction was trained on the dosimetric differences between the IMPT auto-generated plan and the previous photon plan, as well as on the outcomes of the MBS approach. The training process was curated to strongly avoid generating false negatives of IMPT eligible patients. The variables for training compared the differences in xerostomia, dysphagia, and feeding tube dependency for the photon therapy and IMPT plan, and whether or not the MBS approach deems the individual eligible. After training, a three-dimensional decision boundary was generated based on the xerostomia, dysphagia, and feeding tube dependency measurements. The boundary separates pre-selected patients eligible for proton therapy from those that were not pre-selected by the model, and is given by the conditional probability generated from the Gaussian classifier that outlines whether the patient eligible for proton therapy was larger than the decision threshold. Such efficient models provide valuable insight as to whether an individual patient would be selected for proton therapy prior to any other further treatment planning, conserving both time and money.

An alternative model, titled AI-PROTIPP (Artificial Intelligence Predictive Radiation Oncology Treatment Indication to Photons/Protons), features a U-Net architecture and is able to quantitatively assess which treatment method is superior depending on patient factor inputs [13]. The model is able to predict dose distributions for both photon and proton therapy, and use Normal Tissue Complication Probability (NTCP) to determine which therapy would result in the least side effects. This model was employed in clinical cases of patients with oropharyngeal cancer, and resulted in 87.4% accuracy of treatment selection based on the parameters given. AI-PROTIPP effectively uses NTCP data to predict dose distributions, select treatment plans, and reduce the time needed for manual comparisons. Similar to AI-PROTIPP, Chen *et al.* [14] generated two 3D U-Nets to predict photon and proton doses for patients with localized prostate therapy. They selected NTCP models of grade 2 or higher to determine proton therapy partiality. The deep-learning based dose prediction technique returned with 90%-93.5% selection accuracy based on the NTCP model parameters. These studies demonstrate that DL methods can accurately predict dosage and distinguish NTCP differences between photon and proton plans to select the superior modality for the patient [12–14]. Clinically, ML applied to this area has the potential to greatly decrease time spent on plan comparison down to as little as 5 seconds, expediting the start of patient treatment.

Geng *et al.* conducted a multicenter study using a KBP ML model to evaluate and improve radiotherapy plan quality in a clinical trial for non-small-cell lung cancer (NSCLC) [15]. The trial aimed to compare the effectiveness of IMPT versus traditional IMRT, with a focus on reducing toxicity to OARs. The KBP model was utilized to assess the quality of both IMPT and IMRT plans across the various centers, showing that while proton therapy often demonstrated dosimetric advantages, it also had greater variability in plan quality compared to photon therapy. This study highlights the KBP model’s utility in QA, and its potential for enhancing radiotherapy plans in multicenter clinical trials. The research group also plans to publish both photon and proton KBP models for broader use in radiotherapy plan optimization and QA, one of the first for proton therapy.

#### 1.2 Toxicity Prediction and Strategy

Padannayil *et al*. [16] applied a ML algorithm to improve toxicity prediction and to specifically reduce radiation dermatitis for IMPT. Radiation dermatitis is a common side effect of all radiotherapy modalities characterized by skin inflammation and redness. It has been reported that up to 67.4% of nasopharyngeal carcinoma patients experience Grade 2 (medium) radiation dermatitis [17], therefore, finding ways to mitigate its severity is of high importance. The group developed an unsupervised k-means clustering ML algorithm that was trained on previous head and neck cancer patients treated with IMPT. The algorithm created clusters based on dermatitis severity (low, medium, or high grade) and treatment parameters to locate patterns found between the two. By optimizing the distance between clusters, the algorithm was able to group patients according to how likely they are to develop varying grades of radiation dermatitis depending on their treatment parameters. By implementing this model which optimizes where the proton beam is placed, the algorithm is able to lower the skin dose of radiation while maintaining comparable treatment outcomes to traditional photon therapy.

### 2. Simulation & Treatment Planning

The clinical workflow of radiotherapy relies heavily on manual inputs and standardized protocols for simulation and treatment planning, critical components that guarantee safe and effective treatment delivery. While current methodologies are effective, they may be time-consuming, require substantial processing power, and fall susceptible to patient-specific variability. With the incorporation of ML, many of these areas may be automated and optimized to tailor each nuance of treatment planning to the individual patient case.

#### 2.1 Auto-planning and Treatment Plan Optimization

A 3-dimensional cycle generative adversarial network (CycleGAN) was trained on CT and MRI data pairs from a training cohort composed of patients with base-of-skull tumors [18]. The clinical study consisted of 50 patients, split into two non-overlapping cohorts, one for training and one for study purposes. Post-training, synthetic CT (sCT) scans were generated by the algorithm and compared against the initial CT scans where the mean error ranged from 38.65 HU to 65.12 HU. Proton plans with 2 beams each were generated based on the initial CT scans as well as the sCT images, and were compared based on dose-volume histogram endpoints and proton distal range along the beams. Quantitative results showed that the algorithm-generated sCT was in agreement with the ground truth CT; the dosimetric evaluation of the dose-volume histogram endpoints did not have statistically significant differences, and 96% of the proton ranges were clinically accepted. Shafai-Erfani *et al.* showcased an algorithm that is capable of generating clinically-acceptable sCT images and highlighted the potential for MRI-based proton treatment planning, which was subsequently built upon by Zimmerman *et al*. [19].

A DL algorithm capable of generating sCT images independent of MRI sequence data was created in 2022 [19]. 47 meningioma patients treated with pencil beam scanning (PBS) therapy were split into training, validation and test cohorts. MRI sequences in combination with planning CT (pCT) data were used to train a 3D U-Net framework with ResNet-Blocks, a type of CNN block in which the output from one layer is added into a deeper layer. The training outcome was assessed by metric, dosimetric, and spot difference map accuracy in comparison to the original treatment plans. The synthetic dose parameters of the proposed model agreed within 1% of the real-world original plans, and 98% of the spots on the spot difference maps had less than 1 cm difference from the original plan. The novel MRI sequence independent sCT generator created by Zimmerman *et al.* suggests that “the training phase of neural networks can be disengaged from specific MRI acquisition protocols” [19].

Incorporating KBP pipelines improved with ML into commercial treatment planning may decrease inter-plan variability. KBP in the modern day best refers to data-driven approaches in treatment planning, incorporating clinicians’ knowledge into precise algorithms. Post-processing and dose mimicking are critical steps to create clinically acceptable ML-planned IMPT plans. Borderias-Villarroel *et al.* [20] investigated the quality of such plans utilizing four different KBP pipelines. The four models were generated from two sets of conditions; post-processing or no post-processing, isodose-based or commercial RayStation dose mimicking. Using data from 60 oropharyngeal cancer patients, the study trained 11 dose prediction models and results produced comparable target coverage and improved OAR preservation compared to manual plans. However, post-processing appeared to compromise the robustness of the plan. The combination of no post-processing and RayStation dose mimicking produced the IMPT plan with the best robustness with sufficient OAR sparing.

#### 2.2 Target Delineation and Segmentation

Contouring OARs is a laborious, manually-intensive task in the clinical workflow. Studies have shown that contours created by AI are more consistent than those completed manually by oncologists with the same level of NTCP [21]. Applying related technology to the clinical workflow of proton therapy is therefore preferable to ensure patients receive the highest quality treatment plan. Segmentation of the target and OARs utilizing ML has not been explored for proton therapy treatment specifically. At the current time, all published ML algorithms related to contouring in treatment planning pertain to conventional radiation methodologies. Nielsen et al. is the only group to have evaluated a CNN and local AI to contour OARs specifically for future proton therapy [21]. Specific ML models have been developed for adaptive proton therapy purposes, but the planning stage remains underdeveloped.

Van der Veen *et al.* explored how DL models can be implemented in the identification and delineation of OARs in head and neck cancer radiotherapy treatment [22]. While this study does not directly address the applications to proton therapy but rather radiotherapy broadly, the technology has great potential for practical application. Precise delineation of tumors in the head and neck is crucial during treatment planning to ensure that the radiation is sufficiently covering and concentrated on the tumor while sparing surrounding healthy tissue. OAR delineation in the majority of cases is auto-contoured with physician confirmation, but some difficult areas may be delineated manually. The study assesses how a CNN, a type of DL model, can automatically delineate OARs from patient CT or MRI scans used in treatment planning. Using a CNN can significantly decrease the time it takes to delineate the tumor from OAR while simultaneously improving consistency when trained on a large dataset. The authors discuss potential limitations as well: the availability of vast training datasets is still expanding, and the necessity of carefully validating the algorithm’s accuracy remains for successful implementation.

#### 2.3 Dose Prediction, Deposition & Verification

PBS dose calculation often may be inaccurate due to approximations used to work around heterogeneities in tissue, but the result is provided quickly. The doses are calculated using the water equivalent path length along the center of the proton beam, and the medium is treated as infinite and homogeneous. However, tissue is rarely homogenous in clinical reality, leading to range uncertainties and inaccurate modeling of multiple Coulomb scattering. Despite being the current gold standard and extremely accurate, Monte Carlo (MC) dose calculation suffers from slow processing due to tracking each individual particle. To mitigate the tradeoff between accuracy and time when utilizing PBS dose calculation or MC simulations, incorporating ML algorithms may improve the shortcomings of either method.

To boost the accuracy of PBS dose planning while maintaining the short time scale, Wu *et al.* created a DL model that converts a PBS dose to a MC dose using the initial PBS data and the CT images [23]. The model was trained on data from four tumor sites; head and neck, liver, prostate, and lung. The group found that, “Training the model on data from all tumor sites together and using the dose distribution of each individual beam as input yielded the best performance for all four tumor sites” [23]. The average gamma passing rate, a QA measure, was >88% between the converted PBS dose and the MC dose, and the conversion time was less than 4 seconds. Such results demonstrate high accuracy and rapid speed, combining the strengths of both planning techniques. The group reports that the model is able to be applied to new datasets through transfer learning, and may efficiently be added to the clinical workflow of proton therapy treatment planning.

Proton Source Model Commissioning (PSMC) is a critical component of the planning workflow which aims to optimize the match between calculated dose and delivered dose in PBS therapy. Presently, PSMC ensures accurate dose calculation via MC simulations. Nominal energy refers to the average or expected energy value of the protons in the simulation, while the energy spread is the range of energies around the nominal energy. Setting up, or calibrating, the nominal energy and energy spread parameters in the PSMC is difficult as these parameters are not able to be easily solved from an equation. To facilitate these calculations, a CNN known as “PSMC-Net” was developed [24]. PSMC-Net was trained on a range of 33 clinical-level energies, from 75-225 MeV. For each of the 33 energies, a dataset was generated with 15 nominal energies, 10 spreads, and the corresponding 150-calculated depth doses (IDDs). 130 of the data pairs were used for training, 10 for validation, and 10 for testing. Results showed that the gamma pass rate between the MC and measured IDDs was 99% when PSMC-Net was implemented. Without PSMC-Net, the gamma pass rate reduced to ∼54%. This difference in plan quality showcases the significant improvement in accuracy and efficiency of the PSMC process when commissioning a CNN-based proton source model.

Linear energy transfer (LET) refers to the rate at which energy is transferred per unit length. X-rays in photon therapy would be classified as low LET and cause smaller amounts of DNA damage, compared to particles with higher LET, such as alpha particles that subsequently cause greater damage. Protons are advantageous in radiation therapy due to their characteristic Bragg peak, which creates a mixed radiation field of both low LET components further from the tumor and high LET components at the tumor site. MC simulations are utilized as the gold standard to predict yield and LET at a certain depth of the proton treatment plan to assess biological effectiveness. These simulations are difficult to validate experimentally. Gao *et al.* implemented a method to verify LET via an entirely synthetic computational approach for proton stereotactic body radiation therapy (SBRT) [25]. To date, this is the first time a KBP methodology has been adopted in the context of LET distributions. Utilizing the dose distribution map, the DL framework predicts the LET distribution map of the protons to better estimate the relative biological effectiveness (RBE). The framework utilized data from 50 prostate cancer patients receiving proton SBRT, and featured a 5-fold cross-validation method, dividing the patient dataset into five subsets. In each iteration, one subset was used for testing while the remaining four were used for training, ensuring each patient’s data was tested exactly once. The proposed model consists of two sub-networks; two U-net based generators that create a synthetic LET map image from the dose map image, and two discriminations which minimize judgement error. The supervised CycleGAN model demonstrated superior performance to other GAN-based models with a mean absolute error (MAE) of 0.096 ± 0.019 keV *μ*m^−1^. This model may significantly improve the efficacy of proton therapy by improving treatment plan quality through RBE optimization, aided by the provision of highly accurate LET information.

#### 2.4 Range Prediction, Calculation & Verification

An alternative method to estimate proton beam range is the use optical camera systems equipped with scintillators. Scintillators are materials that absorb ionizing radiation, or the energy from charged particles, and convert it into short bursts of visible photons or UV light. Such devices are excellent candidates for dosimetric or range QA measures in radiation therapy. As passive detectors, scintillators provide a non-invasive, safe option for clinical usage. Additionally, the devices are able to provide real-time feedback with high spatial resolution provided by the sensitive ability to capture minute light emissions. They are cost-effective, adaptable to different radiation energies, and generally compatible with optical imaging systems.

Utilizing ML to hybridize dosimetric and range predictions, Ranjith *et al.* developed artificial neural networks (ANN) able to predict six variables related to dosimetric parameters: proton beam spot size in the x-axis, y-axis, major axis, minor axis, and relative x and y-axis positional errors [26]. All of the ANN models utilized a multi-layer perception (MLP) network with one input layer, three hidden layers, and an output layer. Trained on data from 9000 proton spots, all predicted spot size location and relative positional errors were compared with scintillator-measured data as reference. The ANN models resulted in lower prediction errors compared to the scintillators, and demonstrated excellent beam spot size predications as well as positional errors.

Lee *et al.*’s study appears to be one of the first combining scintillators with DL as a QA measure for both range and dose [27]. The group remarks the difficulty previous methods have had with analytically deriving dose distribution from scintillation images manually due to quenching and optical effects. Their study introduces a a 2D residual U-net DL approach to predict beam range and spread-out Bragg peak (SOBP) using a scintillation light distribution (LD) captured in a water phantom. These predictions were reached via 2D map conversion of the LD maps into dose maps. The training and validation set was comprised of 8659 image pairs generated via MC simulations with varying proton beam conditions, and the LD was “reconstructed using photons backpropagated from the aperture as a visual lens” [27]. The model showed high accuracy with beam range and SOBP width resolutions of 0.02 mm and 0.19 mm, respectively, and deviations of less than 0.1 mm and 0.8 mm from reference simulations. Results demonstrated good agreement in gamma analysis, validating the feasibility and accuracy of this DL-based QA method in clinical practice.

#### 2.5 HU-to-SPR Conversion (CT/DECT)

Derived from the nature of proton interactions, stopping power is an essential aspect of treatment planning which measures how quickly protons lose kinetic energy as they travel through a medium. Linear stopping power is proportional to the density of electrons in the absorbing material due to energy loss resulting from Coulombic interactions between the proton and atomic electrons. Linear stopping power is the property that determines where the Bragg peak occurs. The stopping power ratio (SPR) is the ratio of the linear stopping power of the tissue to the stopping power of water [4]. This is an essential parameter in determining how protons will interact with different bodily tissues, such as air, bone, fat, or muscle. To calculate dose distribution, Hounsfield Units (HU) from CT images are converted into SPR values in order to estimate the linear stopping power of tissues, ensuring precise proton dose delivery in the patient. Conventionally, HU-to-SPR conversion is achieved with a calibration curve relating the two units based on known relationships. ML is increasingly being implemented in this clinical step to increase conversion accuracy, reduce systematic errors, and improve time efficiency.

Wang *et al.* present a noise-robust learning-based method to predict RSP maps from DECT images for proton therapy [28]. DECT images may be utilized to derive the RSP maps by obtaining the energy dependence of proton interactions, however the maps are easily compromised due to noise levels and artifacts from physics-based mapping methods. Utilizing ML to reduce the noise when predicting the RSP maps shows great clinical potential. The model developed utilized a residual attention cycle-consistent generative adversarial network (ccGAN), similar to a CycleGAN, to “bring DECT-to-RSP mapping close to a 1-to-1 mapping by introducing an inverse RSP-to-DECT mapping” [28]. In simulation, 20 head-and-neck cancer patients with DECT images were evaluated. A leave-one-out cross-validation strategy evaluated ground truth RSP values which served as learning targets against results from the proposed model. The predicted RSP maps from the model resulted in a mean square error of 2.83% and a MAE less than 3%. Even with additional simulated noise in the DECT datasets, the model maintained a comparably accurate performance, while the traditional physics-based method faced decreased accuracy due to the noise. Regarding DVH metrics, clinical target volumes were less than 0.2 Gy with no statistical significance, with OAR metrics around 1 Gy. The results highlight the excellent accuracy of the predicted RSP maps generated by the ML model and the potential for improving treatment planning and dose calculation.

Translating CT numbers into material properties contributes to uncertainty in dose calculations. A physics-constrained DL-based multimodal imaging (PDMI) framework was proposed to bridge the gap between proton range uncertainty and material density of different tissue types [29]. This model integrated physics, DL, MRI, and DECT to generate accurate mass density maps, and built upon previous work by the same group investigating a similar physics-informed DL framework [30]. The prior network successfully generated accurate mass density and relative SPR maps from DECT images from anthropomorphic adult male, female, and child phantoms with unspecified tissue models. Building upon this work, the new study modeled adipose, brain, muscle, liver, skin, spongiosa, and hydroxyapatite bone in phantoms, and MRI images were taken of each. From the PDMI framework, an empirical model and various residual networks were generated. Parameters included training with and without a physics constraint, and the utilization of MRI and DECT images or solely DECT images. Supervised learning further enhanced the ResNet model trained with a physics constraint compared to the ResNest trial without. Additionally, the ResNet utilizing both MRI and DECT images opposed to solely DECT predicted the densities of each tissue most closely to established literature. As of currently, this novel framework is the first approach to inform DL models with both physics insights and MRI data to derive accurate mass density maps.

A present concern when considering radiotherapy plans for children is to avoid extraneous radiation, including substitution of MR for CT imaging whenever possible. A group affiliated with St. Jude Children’s Research Hospital worked to create synthetic relative proton stopping power (sRPSP) images from MRI sequences utilizing a particular GAN model [31]. Utilizing pCT and MR images from 195 pediatric brain tumor patients, 17 ccGAN models were trained on paired CT-converted RPSP and MRI datasets. T1-weighted, T2-weighted, and FLAIR MRI were tested in 17 total combinations, with or without preprocessing, to cover all optimal training sequences for the models. The intended purpose of the model was to transform the patient MRIs into sRPSP images by learning from the paired CT dataset. To evaluate the performance of the ccGAN and potential for clinical use, the group developed an online QA tool to ensure the safe integration of MR-only proton planning into practice. The generated sRPSP images were converted to sCT, then the QA technique adjusted the sCTs to match a standard reference template created from the training dataset to identify areas where the sCT deviated from the ground-truth CT by >100 HUs. Additionally, a gamma intensity analysis was conducted for similar purposes to analyze the accuracy of the sCT. The group concluded that accurate sRPSP images from T1-weighted and T2-weighted MRI were able to be generated by the ccGAN, with the QA tool highlighting regions of inaccuracy to remove unsuitable sRPSP images from clinical use.

### 3. In-Vivo Monitoring and Adaptivity in Proton Therapy

Continual adaptation and replanning are crucial components of both photon and proton therapy, and demonstrate reductions in NTCP and improved target coverage [32]. The two primary adaptive therapy techniques include offline replanning, which occurs when a new plan is generated between fractions and online replanning, which occurs while the patient is on the treatment table. ML has the potential to benefit both areas of adaptive proton replanning, which currently are time-consuming and potentially require additional irradiation to the patient through repetitive imaging.

#### 3.1 Anatomy Changes

As noted by Wang *et al.*, the sculpting ability benefits of IMPT are countered by increased sensitivity to anatomical variations during and between treatment sessions [33]. As of 2021, anatomic changes were still not effectively accounted for during pediatric treatment plans. Such limitations may result in suboptimal delivery to the tumor or increased dosage to OARs. Adaptive anatomy planning is resource intensive and usually requires additional CT simulation, where in pediatric patients, increased radiation exposure is not preferable. DL may improve this area of clinical workflow by allowing sCTs to be derived from offline on-treatment MRI images to calculate dose delivery based on daily anatomy, as well as flagging cases which may require adaptation.

The group developed a novel CycleGAN model with self-attenuation with abilities to identify pediatric patients that would benefit from adaptive replanning [33]. The goal of creating such a model, opposed to utilizing a conventional CycleGAN, is to assess if the addition of self-attenuation generates more accurate sCTs for children with brain tumors between the ages of 2 and 21. The goal of introducing self-attenuation to a conventional CycleGAN is to enhance boundary delineation of bone-air tissue interfaces, specific to common pediatric brain tumors, and to reduce noise. The training dataset was composed of both CT and T1-weighted MRI images of 125 children with brain tumors. Seven patients between the ages of 2 and 14 that underwent adaptive planning due to anatomy changes discovered via MRI during proton therapy comprised the test dataset. Results were obtained by comparing the MRI taken during proton therapy with the model-generated sCT and the replanning CT (ground truth). The HU MAE with the self-attenuation CycleGAN was 65.3 ± 13.9 versus 88.9 ± 19.3 for the conventional CycleGAN, demonstrating improved accuracy by the proposed model. The self-attenuated model also demonstrated improved gamma passing rates and appropriately triggered plan adaptation in all test patients.

Similar to the study utilizing a CNN to contour OARs in conventional radiation planning [22], Elmahdy *et al.* utilized a CNN to automatically segment the bladder from CT scans of IMPT prostate cancer patients online [34]. This study focuses on online adaptive IMPT, where the patient’s anatomy is continuously tracked to generate real-time treatment adjustments in the case of observed changes. In the case of prostate cancer, the goal of this algorithm was to monitor changes in bladder or prostate shape or location to minimize OAR dosage. Particular attention was paid to accurate segmentation of the bladder and adjacent structures (prostate, seminal vesicles, and lymph nodes). The group acknowledges commercially available automatic recontouring applications for adaptive therapy, but reaffirms their current status as a black box for clinicians.

The proposed CNN is able to predict deformation vector fields which are required for accurate contour propagation by detecting spatial transformations between the reference CT image and the real-time treatment imaging [34]. The CNN was trained on manually-delineated images and corresponding deformation vector fields of 20 prostate cancer patients. As the network makes the continuous comparison between the two images, this enables adaptations in the treatment plan to be generated based on the patient’s anatomical variations during the treatment session. Their results found that the combined CNN and image registration technique improved the target delineation accuracy significantly with a dice similarity coefficient of 88% compared to manual attempts. The CNN-propagated contours met dose coverage constraints in 86%, 91%, and 99% of cases for the prostate, seminal vesicles, and lymph nodes, respectively. The group reported that 80% of the automatically generated treatment plans were directly usable without manual correction, improving clinical efficiency and potentially reducing adverse side effects by sparing healthy tissue.

#### 3.2 Range

Protoacoustic signals in proton therapy are acoustic waves generated when protons interact with tissues. According to the Bragg peak, protons suddenly and rapidly deposit energy at a certain depth, causing a local transient increase in temperature and pressure [35]. This minute expansion generates protoacoustic signals, which may be detected with ultrasound sensors. The clinical relevance relates to recreation of the Bragg peak and real-time monitoring of proton beam range, ensuring that the beam is depositing at the intended target depth. Protoacoustic signal denoising is quickly gaining traction in the field of proton therapy research, and ML techniques are readily being incorporated.

While the protoacoustic technique is able to determine the Bragg peak location in vivo, it requires a large dose delivery in order to achieve an adequate signal-to-noise (SNR) ratio and high number of signaling average (NSA). Such large doses are not suitable for clinical use. A novel DL model demonstrated enhancement of the SNR of protoacoustic measures as well as improving Bragg peak location identification accuracy in range verification scenarios [36]. Three accelerometers were placed on cylindrical polyethylene phantoms to record protoacoustic signals during simulated treatment. Device-specific stack autoencoder (SAE) denoising models were trained to denoise low NSA signals. The supervised SAE outperformed the unsupervised SAE in Bragg peak range verification. Clinically, this model shows promise to decrease unnecessary dosage in the patient by accurately measuring the Bragg peak in vivo.

Expanding upon the range of applications of ML in online range monitoring, Jiang *et al*. proposed a novel architecture capable of denoising acoustic, protoacoustic, and electroacoustic signals both quantitatively and qualitatively [37]. Radiation-induced acoustic imaging generally requires a substantial number of recorded frames to achieve a satisfactory dose deposition average, exposing the patient to increased dose. The proposed model is a general deep inception convolution neural network, or a GDI-CNN. The model features radiation-induced acoustic signal denoising capabilities, which results in a reduced number of frames required for averaging. The group details that “the network employs convolutions with multiple dilations in each inception block, allowing it to encode and decode signal features with varying temporal characteristics” [37]. Such characteristics expand the reach of the network’s abilities to different radiation sources. Compared to prior inception networks, this study implements convolutions with different dilations, rather than different filter sizes, resulting in increased inception field size and decreased computational assumptions. The inputs of the model were the few-averaged noisy signals, which in turn outputs high-SNR denoised signals. For the protoacoustic radiation therapy trial, four of the five data were employed for model training, with the remainder used for validation. The performance of the GDI-CNN was assessed in comparison with experimental data of protoacoustic signals; it realized proton range accuracy parallel to 1500-frame-average results with only 20-frame-average measurements inputted. These results improved range verification frequency from 0.5 to 37.5 Hz in proton therapy. Additionally, it demonstrated lower mean-squared errors and higher peak-SNR, improving the reach of real-time proton therapy monitoring.

#### 3.3 Dosimetry

Protoacoustic imaging specifically focuses on generating images based on the acoustic signals generated by protons. This specific application of protoacoustic signaling is beneficial as it offers in vivo 3D dose verification; however, it is limited by the narrow angle of the ultrasound transducer. Jiang’s group developed a DL model featuring a deep cascaded CNN (DC-CNN) to improve proton-acoustic image reconstruction using proton-acoustic signals detected by a matrix array [38]. The framework was validated on data from 81 prostate cancer patients’ IMPT therapy plans, and dosage was calculated with RayStation. A matrix ultrasound array was simulated near patient treatment site to measure radiofrequency signals during dose delivery. To address how realistic the simulation may be, tissue heterogeneity and attenuation were considered by adding noise to the signals. The proposed model was trained on 204 samples from 69 patients and tested on 26 samples from 12 separate patients. 3D pressures and dose maps were generated, and qualitatively and quantitatively compared with the ground truth. The results showed that the proposed DC-CNN reconstructed high-quality 3D pressure images from the proton-acoustic images, potentially enabling 3D dose verification during treatment.

Pastor-Serrano *et al.* propose DoTA, a DL based millisecond speed dose calculation algorithm capable of accurately predicting pencil beam proton dose depositions [39]. The two current physics-based tools to calculate proton doses are analytical pencil beam algorithms (PBA) and MC simulations, which respectively feature better speed or precision. MC methods and PBAs are currently unable to synthesize the particle transport simulations in sub-second times necessary for next generation real-time adaptive radiotherapy. The importance of accurate particle transport calculations results from IMPT requiring MC simulations and PBAs to compute the spatial distribution of physical dose delivered by the many individual protons. Based upon previous long short-term memory (LSTM) networks [40, 41], the proposed algorithm sequentially calculates proton pencil beam dose distributions from relative SPR slices, and does not require a separate model per beam energy [39]. DoTA takes a novel approach by featuring an attention-based transformer backbone, which dispenses of convolutions and connects the encoder and decoder. This architecture is simpler than many other transduction models, which accounts for the additional speed that may benefit adaptive replanning.

With the parametric DoTA model, dose distributions from individual proton beamlets are able to be predicted from patient geometries (x) and beam energies (ɛ̝) with remarkable speed. The essential concept behind the algorithm is that it captures particle transport physics from provided data and learns the appropriate function y(θ)= f(x, ɛ̝) through a series of artificial neural networks with parameters (θ). Performance was based on speed and accuracy comparison to standard clinically used methods, such as PBA or MC ground truth dose distributions. In summary, DoTA predicted dose distributions from single pencil beams 100 times quicker than widely used PBAs. The distributions yielded a gamma pass rate of 99.37 ± 1.17, close to standard MC accuracy in a fraction of the computational time. DoTA outperformed previous PBA and MC approaches with a 10% improvement in gamma pass rate, and features speed close to commercial GPU MC methods, critical for adaptive replanning events.

### 4. Future directions

Significant advances have been recently made incorporating ML into the proton therapy clinical workflow. However, this is still an emerging area of investigation gaining momentum as proton therapy becomes more commonplace. While great strides have been made in method applicable to all radiotherapy generally, proton therapy-specific models have been less frequently published. Areas of priority for future development are discussed in the following section.

#### 4.1 Quality Assurance

At present, ML-assisted QA methods are sparse in proton therapy. QA is laborious in all high-precision radiation therapy modalities, and is comprises management of beam delivery mechanisms, beam parameters, and instrumentation. Currently, ML algorithms have been applied to improving gamma pass rates and predicting beam data in proton therapy, but no publications directly address ML for proton QA. Prerequisites for a quality plan, such as accurate delineation of the target and OARs, is beyond the scope of most QA procedures [15]. According to the AAPM Task Group 224, QA procedures have been outlined for three proton therapy techniques: scattering, uniform scanning, and PBS [42]. This is an area of work where ML may be incorporated to improve the accuracy, efficiency, and consistency in QA. Ono *et al.* reported on the applications of AI for QA in radiotherapy as of 2024, and proton therapy is absent [43]. Current focus areas include feature extraction and selection, mechanical hardware setup QA, and gamma passing rate calculations. With the recently developed QA procedure outlines by Task Group 224 [42], ML may now be addressed as it has been with various other radiotherapy techniques. Connecting back to ML for in-vivo monitoring, an underdeveloped area, projects can continue to be pursued to further improve real-time treatment monitoring by analyzing live delivery of dosage. Additionally, AI may be harnessed to provide streamlined QA workflows by automating manual routine tasks, such as data entry or documentation.

#### 4.2 Cost Calculation

A large barrier to proton therapy treatment is cost. Proton therapy is expensive and may be inaccessible to patients not geographically located near a treatment center. By training on patient geographic and socioeconomic data and associated treatment outcomes, ML could be leveraged to improve equity or personalize the costs of proton treatment. From an institutional perspective, these same data could be used to assess opportunities for growth. Verma *et al.* investigated the health economics of proton therapy in terms of sustainability and cost-effectiveness [44]. As this study was conducted purely from a health economics perspective, ML was not utilized. A significant concern brought to attention is the inherent inaccuracy of cost-effectiveness analyses due to the inability to account for all aspects of operation, such as electricity and maintenance, beam delivery time, and number of patients treated on a given day. The group reported that Markov and MC modeling have been utilized to address cost tabulations and comparisons, but all modeling studies have limitations due to probabilistic assumptions. As of 2016 when the study was conducted, the group reports that proton therapy for the routine treatment of breast cancer was not shown to be cost-effective. ML may greatly improve the accuracy and efficiency of cost-effectiveness analyses by reliably accounting for additional factors otherwise neglected through manual calculation [44]. Additionally, as demonstrated in previously discussed studies, ML is able to improve MC simulations in various aspects [23, 39, 45]. However, such an approach has not yet been accounted for in proton therapy economic analysis.

#### 4.3 Outcome Studies

Prognostic research is sparse in the context of both proton therapy and ML. The recent nature of proton therapy compared to the history of data available for conventional photon therapy is a primary limiting factor. ML has been utilized to predict toxicity and side effects [16, 46, 47], but not overall treatment prognostics in proton therapy. The integration of DL radiomics and circulating tumor cell counts has demonstrated improved accuracy in predicting the risk of reoccurrence for NSCLC patients treated with stereotactic body radiotherapy (SBRT). Jiao *et al.* accomplished this feat by building DL models on clinical measures and CT data from 421 NSCLC patients [48]. The models were able to not only predict recurrence risks, but were able to stratify patients into specific outcome subgroups based on the CTC counts. The proposed model achieved a concordance index of 0.880. It is promising for such a novel methodology to be applied to proton therapy subsequently, gleaning valuable predictions for physicians to consider throughout the treatment process. In particular, such a technique may serve beneficial in adaptive replanning.

#### 4.4 Challenges

The recent nature of proton therapy compared to the history of data available for conventional photon therapy is a limiting factor in the incorporation of ML. There are many large, high-quality datasets available for conventional radiation therapy, but few for proton therapy. This limits most training and testing data for proton therapy to clinical work, as deidentified and declassified patient datasets are not as common. Additional challenge arises from the variation in the nature of the physics. Proton therapy harnesses the complex power of the Bragg peak to provide targeted therapy with decreased side effects. However, with such benefit comes difficulty in applying existing ML algorithms used in conventional radiotherapy to proton therapy. ML applications are common in other areas of radiation therapy, but the challenge in adapting and applying them to proton therapy is inherent to the predicate physics and hardware differences.

## Discussion

Many variations and applications of machine learning (ML) have appeared in the proton therapy workflow over the past five years with promise of improving efficiency, accuracy, and efficacy of treatment. U-Net architectures are prevalent in the patient pre-screening process, and convolutional neural networks (CNNs) and CycleGANs frequent dose and range prediction in treatment planning. For adaptive monitoring, advanced deep learning architectures such as general deep inception convolution neural networks (GDI-CNNs) and deep cascaded CNNs (DC-CNNs) improve real-time dose verification and range monitoring. However, certain areas such as target segmentation in treatment planning, cost analysis, and quality assurance measures lack notable ML contributions and are scopes of future interest. In conclusion, with the increased clinical interest in proton therapy, ML algorithms have been incorporated into both the clinical and research workflows to facilitate treatment and discovery.

**Table 1:**
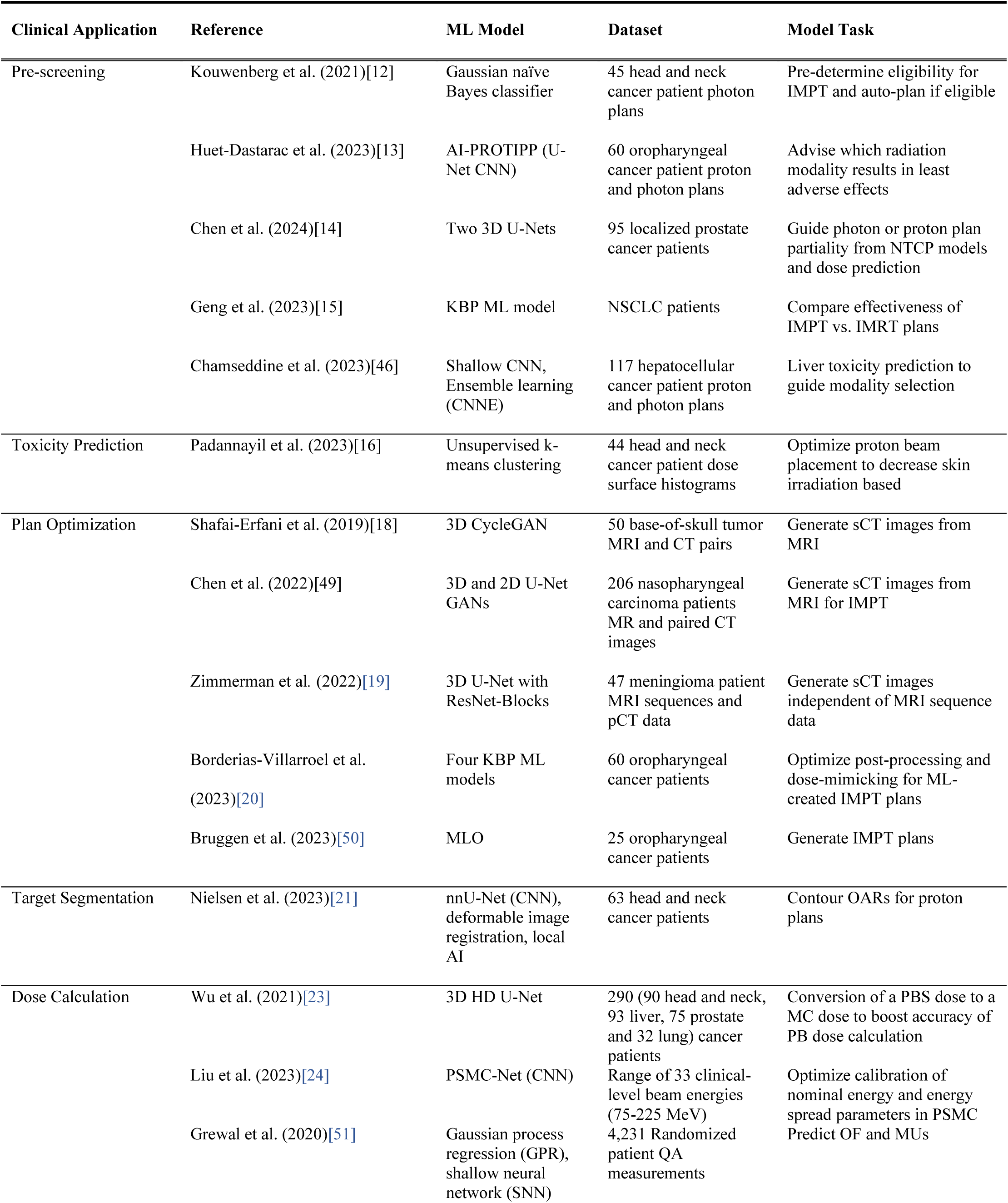
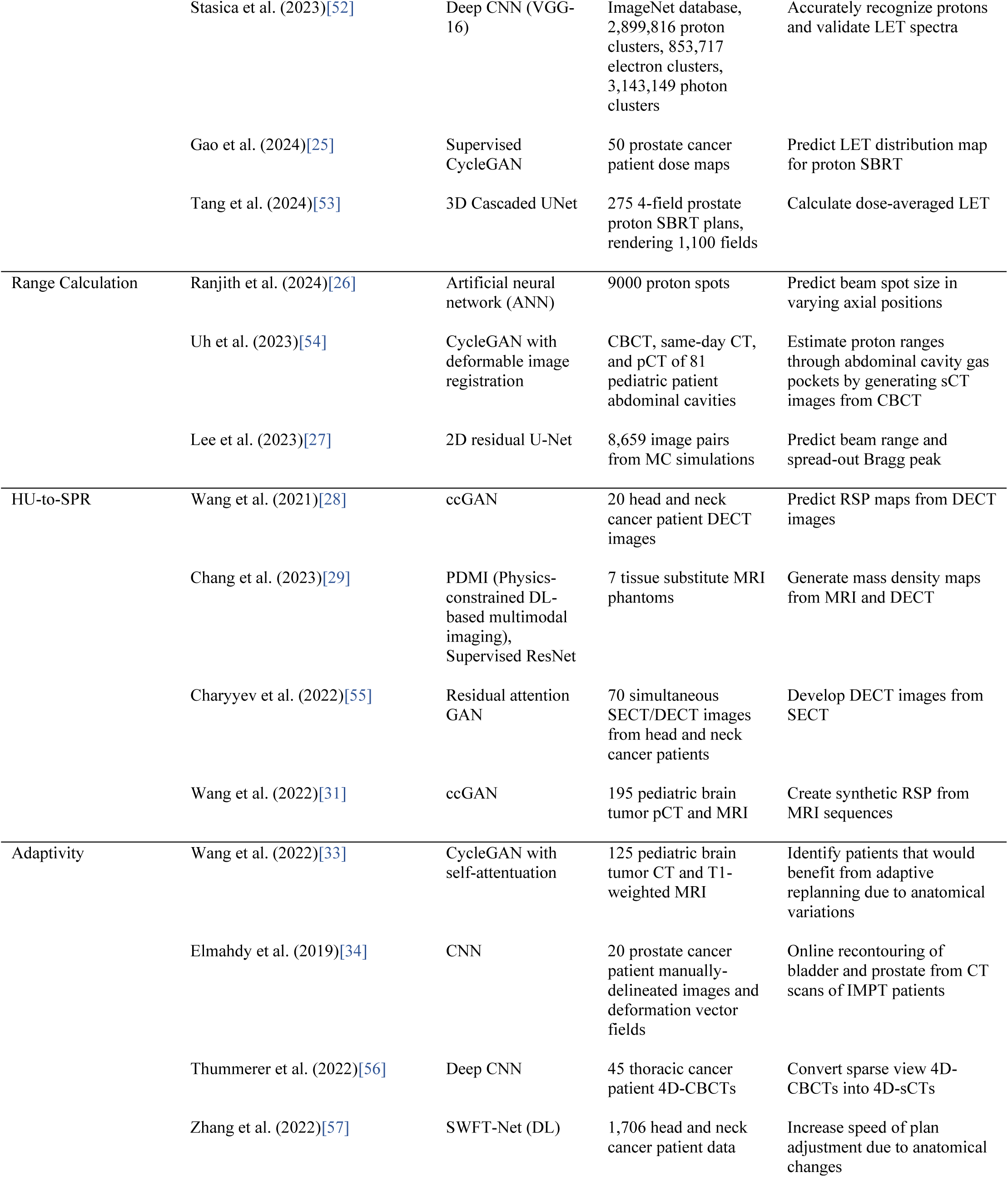
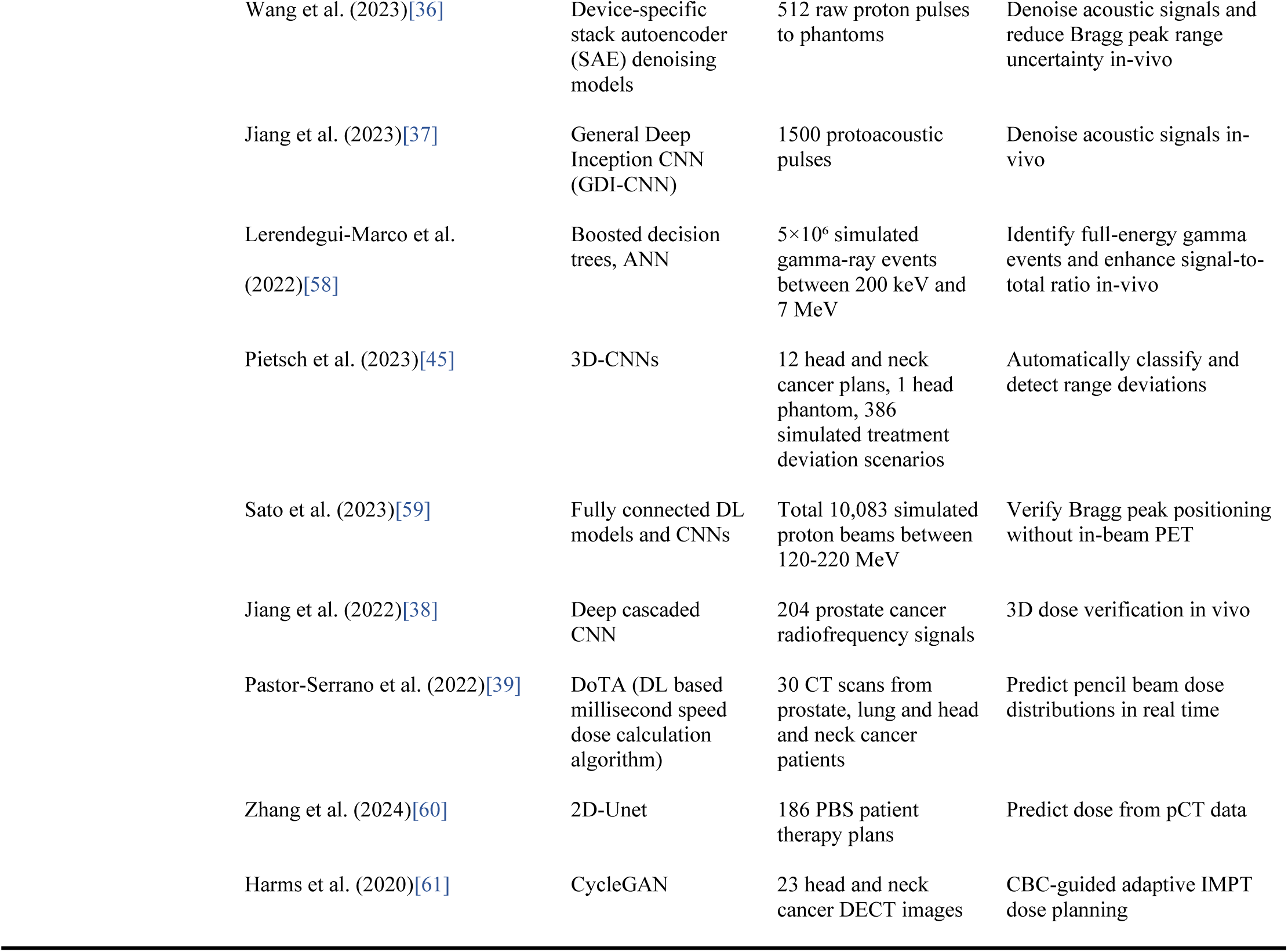
Datasheet summary of current ML applications and associated data sources at various stages of proton therapy clinical treatment.

| Clinical Application | Reference | ML Model | Dataset | Model Task |
| --- | --- | --- | --- | --- |
| Pre-screening | Kouwenberg et al. (2021)[12] | Gaussian naïve Bayes classifier | 45 head and neck cancer patient photon plans | Pre-determine eligibility for IMPT and auto-plan if eligible |
|  | Huet-Dastarac et al. (2023)[13] | AI-PROTIPP (U-Net CNN) | 60 oropharyngeal cancer patient proton and photon plans | Advise which radiation modality results in least adverse effects |
|  | Chen et al. (2024)[14] | Two 3D U-Nets | 95 localized prostate cancer patients | Guide photon or proton plan partiality from NTCP models and dose prediction |
|  | Geng et al. (2023)[15] | KBP ML model | NSCLC patients | Compare effectiveness of IMPT vs. IMRT plans |
|  | Chamseddine et al. (2023)[46] | Shallow CNN, Ensemble learning (CNNE) | 117 hepatocellular cancer patient proton and photon plans | Liver toxicity prediction to guide modality selection |
| Toxicity Prediction | Padannayil et al. (2023)[16] | Unsupervised k-means clustering | 44 head and neck cancer patient dose surface histograms | Optimize proton beam placement to decrease skin irradiation based |
| Plan Optimization | Shafai-Erfani et al. (2019)[18] | 3D CycleGAN | 50 base-of-skull tumor MRI and CT pairs | Generate sCT images from MRI |
|  | Chen et al. (2022)[49] | 3D and 2D U-Net GANs | 206 nasopharyngeal carcinoma patients MR and paired CT images | Generate sCT images from MRI for IMPT |
|  | Zimmerman et al. (2022)[19] | 3D U-Net with ResNet-Blocks | 47 meningioma patient MRI sequences and pCT data | Generate sCT images independent of MRI sequence data |
|  | Borderias-Villarroel et al. (2023)[20] | Four KBP ML models | 60 oropharyngeal cancer patients | Optimize post-processing and dose-mimicking for ML-created IMPT plans |
|  | Bruggen et al. (2023)[50] | MLO | 25 oropharyngeal cancer patients | Generate IMPT plans |
| Target Segmentation | Nielsen et al. (2023)[21] | nnU-Net (CNN), deformable image registration, local AI | 63 head and neck cancer patients | Contour OARs for proton plans |
| Dose Calculation | Wu et al. (2021)[23] | 3D HD U-Net | 290 (90 head and neck, 93 liver, 75 prostate and 32 lung) cancer patients | Conversion of a PBS dose to a MC dose to boost accuracy of PB dose calculation |
|  | Liu et al. (2023)[24] | PSMC-Net (CNN) | Range of 33 clinical-level beam energies (75-225 MeV) | Optimize calibration of nominal energy and energy spread parameters in PSMC |
|  | Grewal et al. (2020)[51] | Gaussian process regression (GPR), shallow neural network (SNN) | 4,231 Randomized patient QA measurements | Predict OF and MUs |
|  | Stasica et al. (2023)[52] | Deep CNN (VGG-16) | ImageNet database, 2,899,816 proton clusters, 853,717 electron clusters, 3,143,149 photon clusters | Accurately recognize protons and validate LET spectra |
|  | Gao et al. (2024)[25] | Supervised CycleGAN | 50 prostate cancer patient dose maps | Predict LET distribution map for proton SBRT |
|  | Tang et al. (2024)[53] | 3D Cascaded UNet | 275 4-field prostate proton SBRT plans, rendering 1,100 fields | Calculate dose-averaged LET |
| Range Calculation | Ranjith et al. (2024)[26] | Artificial neural network (ANN) | 9000 proton spots | Predict beam spot size in varying axial positions |
|  | Uh et al. (2023)[54] | CycleGAN with deformable image registration | CBCT, same-day CT, and pCT of 81 pediatric patient abdominal cavities | Estimate proton ranges through abdominal cavity gas pockets by generating sCT images from CBCT |
|  | Lee et al. (2023)[27] | 2D residual U-Net | 8,659 image pairs from MC simulations | Predict beam range and spread-out Bragg peak |
| HU-to-SPR | Wang et al. (2021)[28] | ccGAN | 20 head and neck cancer patient DECT images | Predict RSP maps from DECT images |
|  | Chang et al. (2023)[29] | PDMI (Physics-constrained DL-based multimodal imaging), Supervised ResNet | 7 tissue substitute MRI phantoms | Generate mass density maps from MRI and DECT |
|  | Charyyev et al. (2022)[55] | Residual attention GAN | 70 simultaneous SECT/DECT images from head and neck cancer patients | Develop DECT images from SECT |
|  | Wang et al. (2022)[31] | ccGAN | 195 pediatric brain tumor pCT and MRI | Create synthetic RSP from MRI sequences |
| Adaptivity | Wang et al. (2022)[33] | CycleGAN with self-attention | 125 pediatric brain tumor CT and T1-weighted MRI | Identify patients that would benefit from adaptive replanning due to anatomical variations |
|  | Elmahdy et al. (2019)[34] | CNN | 20 prostate cancer patient manually-delineated images and deformation vector fields | Online recontouring of bladder and prostate from CT scans of IMPT patients |
|  | Thummerer et al. (2022)[56] | Deep CNN | 45 thoracic cancer patient 4D-CBCTs | Convert sparse view 4D-CBCTs into 4D-sCTs |
|  | Zhang et al. (2022)[57] | SWFT-Net (DL) | 1,706 head and neck cancer patient data | Increase speed of plan adjustment due to anatomical changes |

|  |  |  |  |
| --- | --- | --- | --- |
| Wang et al. (2023)[36] | Device-specific stack autoencoder (SAE) denoising models | 512 raw proton pulses to phantoms | Denoise acoustic signals and reduce Bragg peak range uncertainty in-vivo |
| Jiang et al. (2023)[37] | General Deep Inception CNN (GDI-CNN) | 1500 protoacoustic pulses | Denoise acoustic signals in-vivo |
| Lerendegui-Marco et al. (2022)[58] | Boosted decision trees, ANN | $5 \times 10^6$ simulated gamma-ray events between 200 keV and 7 MeV | Identify full-energy gamma events and enhance signal-to-total ratio in-vivo |
| Pietsch et al. (2023)[45] | 3D-CNNs | 12 head and neck cancer plans, 1 head phantom, 386 simulated treatment deviation scenarios | Automatically classify and detect range deviations |
| Sato et al. (2023)[59] | Fully connected DL models and CNNs | Total 10,083 simulated proton beams between 120-220 MeV | Verify Bragg peak positioning without in-beam PET |
| Jiang et al. (2022)[38] | Deep cascaded CNN | 204 prostate cancer radiofrequency signals | 3D dose verification in vivo |
| Pastor-Serrano et al. (2022)[39] | DoTA (DL based millisecond speed dose calculation algorithm) | 30 CT scans from prostate, lung and head and neck cancer patients | Predict pencil beam dose distributions in real time |
| Zhang et al. (2024)[60] | 2D-Unet | 186 PBS patient therapy plans | Predict dose from pCT data |
| Harms et al. (2020)[61] | CycleGAN | 23 head and neck cancer DECT images | CBC-guided adaptive IMPT dose planning |

## Supporting information

Funding Information

## Data Availability

All data and studies referenced are publicly available in the cited sources.

